# Investigating Neural Processing of Color in Normal and Impaired Vision

**DOI:** 10.1101/2024.12.22.24319498

**Authors:** Andriani Rina

## Abstract

This study examines cortical responses to chromatic and luminance stimuli in individuals with normal trichromatic vision, Daltonism, and achromatopsia. Functional magnetic resonance imaging (fMRI) data were collected using stimuli modeled after Wade et al. (2008) to evaluate the differential activation of visual cortical areas. In normal trichromats, hV4 demonstrated the highest chromatic sensitivity, while ventral areas showed stronger responses to color compared to dorsal regions. In Daltonic and achromatopsia participants, cortical activation was observed under combined chromatic and luminance conditions; however, no significant color-specific activity was detected, even in hV4.

This work establishes a baseline for understanding cortical responses in color vision deficiencies and preceded gene therapy studies in the same achromatopsia patients (Fischer et al., 2020; Seitz et al., 2022). These findings contribute to ongoing research into neural plasticity and targeted therapeutic interventions.

## Introduction

The human visual system processes information about color and luminance through distinct neural pathways. In trichromatic vision, three cone types—short-wavelength (S, blue-sensitive), medium-wavelength (M, green-sensitive), and long-wavelength (L, red-sensitive)—work in concert to provide a full spectrum of color perception. These signals are processed primarily in the ventral visual stream, including V1, V2v, V3v, and hV4, which are specialized for chromatic processing. In contrast, dorsal areas such as V3d and V3A are more responsive to luminance.

Daltonism, a common form of color vision deficiency, disrupts this balance due to mutations affecting M or L cones. This condition impairs chromatic pathways and forces reliance on luminance-based cues for color discrimination, a phenomenon known as metamerism. Achromatopsia represents a more profound deficit, as individuals with this condition lack functional cones altogether, relying solely on rod-mediated vision. This rod-dominated visual processing results in complete color blindness and severely reduced visual acuity.

The study of Daltonism and achromatopsia provides critical insights into the relationship between retinal input and cortical processing, offering a window into the adaptability and plasticity of the visual system. Daltonism highlights how the brain compensates for deficits at the retinal level by utilizing luminance-based mechanisms to maintain functional vision. Achromatopsia, in contrast, allows for the investigation of cortical reorganization in the complete absence of chromatic input, shedding light on how the visual cortex adapts to significant sensory deficits.

With the advent of gene therapy targeting CNGA3 mutations, such as those reported by Fischer et al. (2020), understanding the pre-treatment neural activity in these patients becomes crucial for evaluating therapeutic outcomes. These therapies, which aim to restore cone function, hold great promise for improving vision in achromatopsia patients. Recent studies, including Seitz et al. (2022), have highlighted the neural plasticity underlying these therapeutic effects, emphasizing the importance of baseline studies like this one for assessing the efficacy of emerging treatments.

By studying these populations, this work not only advances our understanding of neural mechanisms underlying color vision deficiencies but also has broader implications for sensory neuroplasticity. Furthermore, the findings may inform the development of rehabilitation strategies for other acquired visual impairments, providing a translational bridge between basic neuroscience and clinical applications.

This study examines neural activity in the visual cortex of trichromatic, Daltonic, and achromatopsia participants using functional magnetic resonance imaging (fMRI). By presenting luminance-only, chromatic-only, and combined luminance-chromatic stimuli, we hypothesized that a. Ventral visual regions would exhibit reduced chromatic responses in color-deficient participants, particularly in achromatopsia cases, b. Daltonic individuals would rely on luminance-based compensatory mechanisms for chromatic discrimination and c. Achromatopsia participants would demonstrate reduced cortical activity across all conditions, reflecting their rod-dominated visual processing.

These experiments served as a baseline investigation for exploring cortical activation in achromatopsia patients.. By employing tailored fMRI paradigms inspired by Wade et al. (2008), the study provides a comprehensive analysis of cortical activity, addressing gaps in the literature and offering a platform for future investigations into treatment efficacy and neurorehabilitation.

## Methods

### Participants

Seven participants were included in this study: four normal trichromatic individuals (two males and two females, aged 22–26) and three colorblind participants (two females and one male, aged 23–38). One of the colorblind participants had Daltonism and lacked functional L-cones, while the other two were diagnosed with achromatopsia caused by CNGA3 mutation. These mutations resulted in complete rod-mediated vision under mesopic conditions due to the non-functional cones. Ethical approval was granted by the Medical Faculty of the University of Tübingen, and all participants provided written informed consent. Participants were compensated for their time and informed of their right to withdraw at any point without explanation.

### MRI Data Acquisition

MRI data were collected at the Max Planck Institute for Biological Cybernetics, Tübingen, Germany, using a 3.0T Siemens Prisma scanner equipped with a 64-channel posterior head coil. High-resolution T1-weighted anatomical images were acquired using a magnetization-prepared rapid acquisition gradient echo sequence (ADNI protocol) with the following parameters: voxel size = 1 × 1 × 1 mm³, TR = 2000 ms, TE = 3.06 ms, TI = 1100 ms, flip angle = 9°, and matrix size = 256 × 256. Functional scans were performed using a gradient-echo EPI sequence optimized for occipital cortex activation. Each functional scan consisted of 29 slices with a voxel size of 3 × 3 × 3 mm³, TR = 2000 ms, TE = 35 ms, flip angle = 90°, and matrix size = 64 × 64. Each run comprised 181 volumes, with up to five runs collected per participant. The slices covered the occipital lobe and extended into the cerebellum to ensure full coverage of visual processing areas.

### Experimental Design

The study consisted of two experiments designed to assess neural responses to visual stimuli: retinotopy mapping and a color/luminance experiment. Stimuli for both experiments were generated using Psychtoolbox-3 (PTB-3) in MATLAB and presented on a gamma-corrected screen measuring 41 × 24 cm, with a refresh rate of 60 Hz and a resolution of 1280 × 1024 pixels. Gamma correction was performed using a Konica Minolta CS-100SA spectroradiometer to ensure linearized luminance across all RGB channels. Participants viewed the stimuli through a mirror mounted on the MRI head coil, positioned 12 cm from their eyes and 61 cm from the screen.

### Stimuli and Task

#### Retinotopy Mapping

During the experiment, participants viewed high-contrast, moving square-checkerboard bars. These bars traveled seamlessly in eight different directions (e.g., 0°, 45°, 90°, etc.), displayed within a circular aperture with an 11.25° radius centered on the fixation point. Each bar had a width of 1.875° and advanced in steps of 0.9375°, which is half its width, synchronized with the acquisition of each image volume (TR = 2 seconds). The visual stimuli were created using MATLAB, with the assistance of Psychtoolbox (version 3) and the Vistadisp open-source toolbox.

#### Color/Luminance Experiment

The color/luminance experiment assessed neural responses to luminance-only, chromatic-only, and combined luminance and chromatic contrasts. The design of the stimuli was based on previous work by Wade et. al (2008), with modifications to suit the study’s specific objectives. Stimuli were presented as a 22 × 22 checkerboard grid, with each square subtending 1° of visual angle (total grid size: 22° × 22°). Colors and luminance of the squares were refreshed every second (1 Hz) to provide temporal and spatial contrast. Blank intervals (baseline) were interleaved between stimulus conditions to allow hemodynamic activity to return to baseline (Fig.1).

**Figure 1:**
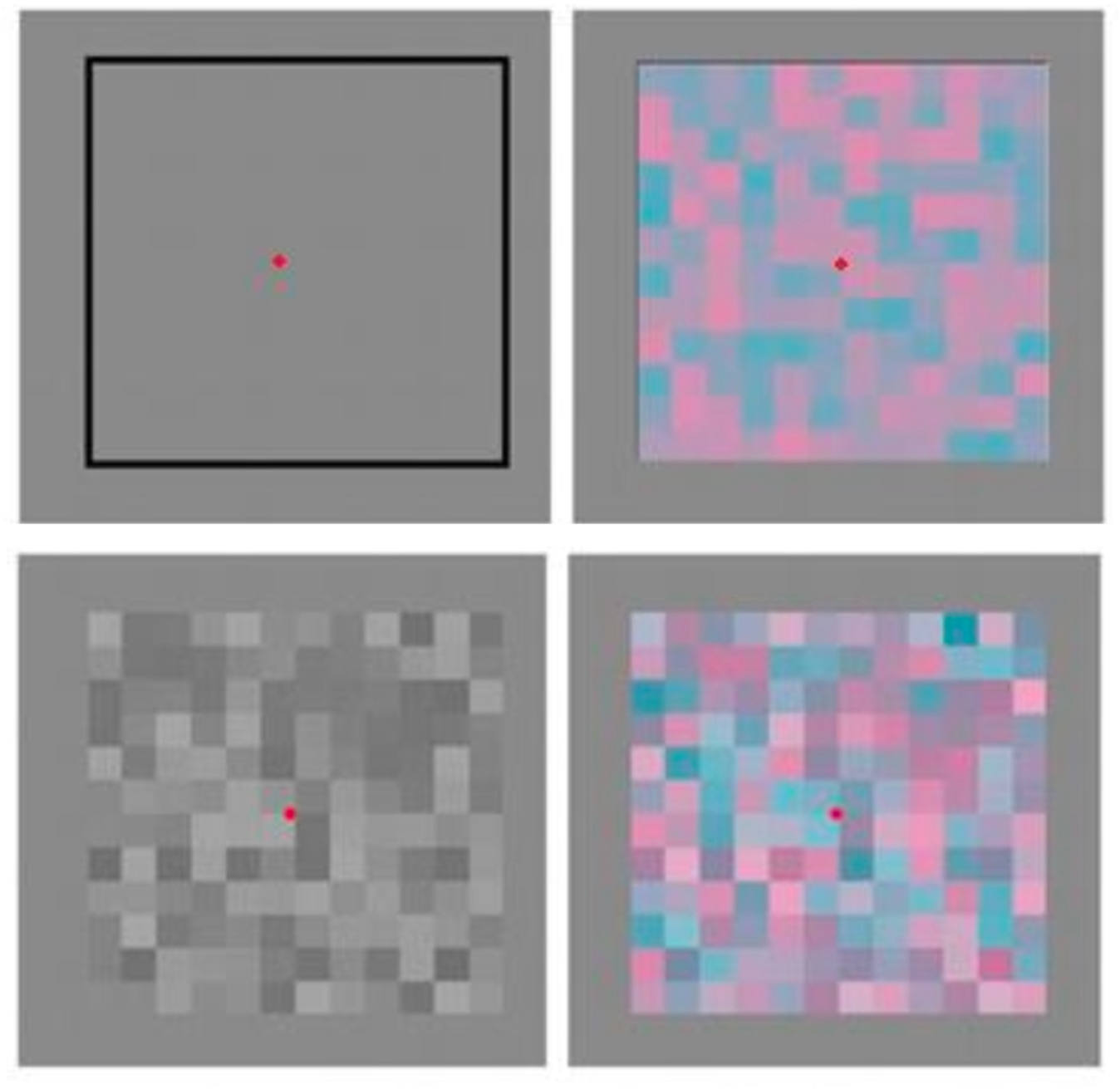
Stimuli Used in the Experiment. The figure displays the stimuli used during the experiment. *Top left:* The baseline/intertrial design. *Top right*: A red/green chromatic stimulus with a maximum 6% (L-M)-cone contrast, without added luminance. *Bottom left*: An achromatic stimulus representing luminance (L+M+S). *Bottom right*: A chromatic stimulus with added luminance ((L+M+S) + (L-M)).

Four conditions were presented:

1. **Luminance Contrast Only (L+M+S):** Squares varied in brightness, with random achromatic contrasts up to **24%**. All three cone types (L, M, and S) were modulated equally, ensuring the stimuli were purely luminance-based. Brighter squares increased excitation proportionally (e.g., LMS = [0.62, 0.62, 0.62]), while darker squares decreased excitation proportionally (e.g., LMS = [0.38, 0.38, 0.38]).
2. **Chromatic Contrast Only (Red-Green; L-M):** Chromatic contrasts of up to **6%** were created by modulating L (long-wavelength, red-sensitive) and M (medium-wavelength, green-sensitive) cones in opposite directions, while maintaining constant S cone excitation and overall luminance. For example, increasing L cone excitation and decreasing M cone excitation produced redder squares (e.g., LMS = [0.53, 0.47, 0.5]), while the reverse produced greener squares (e.g., LMS = [0.47, 0.53, 0.5]).
3. **Chromatic Contrast Only (Blue-Yellow; L+M-S):** Chromatic contrasts were achieved by modulating L+M cones relative to the S cone while holding overall luminance constant. For example, increasing L+M excitation and reducing S excitation produced yellow squares (e.g., LMS = [0.56, 0.56, 0.38]), while the reverse produced blue squares (e.g., LMS = [0.44, 0.44, 0.62]).
4. **Combined Luminance and Chromatic Contrast (L+M+S + L-M):** This condition combined brightness and hue variations, where luminance (L+M+S) and chromatic (L-M) contrasts were simultaneously modulated. For example, adding L+M+S = [0.6, 0.6, 0.6] to L-M = [0.53, 0.47, 0.5] resulted in LMS = [0.63, 0.57, 0.6], appearing brighter and redder.

### Achromatopsia-Specific Adjustments (psychopysical test)

For participants with achromatopsia, stimuli were calibrated through a pre-experiment psychophysical task. In this task, a circle divided into red and green semicircles was displayed. The contrast of the red semicircle was kept constant at 6%, while participants used an MRI-compatible device to adjust the luminance of the green semicircle. They continued making adjustments until they perceived the two halves as having equal brightness and reported their final decision. The green luminance values ranged from 1 to 1.5, changing in steps of 0.025, with 1 representing the luminance level that matches the red semicircle’s luminance as perceived by normal trichromatic vision and 1.5 corresponding to an achromatic luminance level (Fig.2).

**Figure 2:**
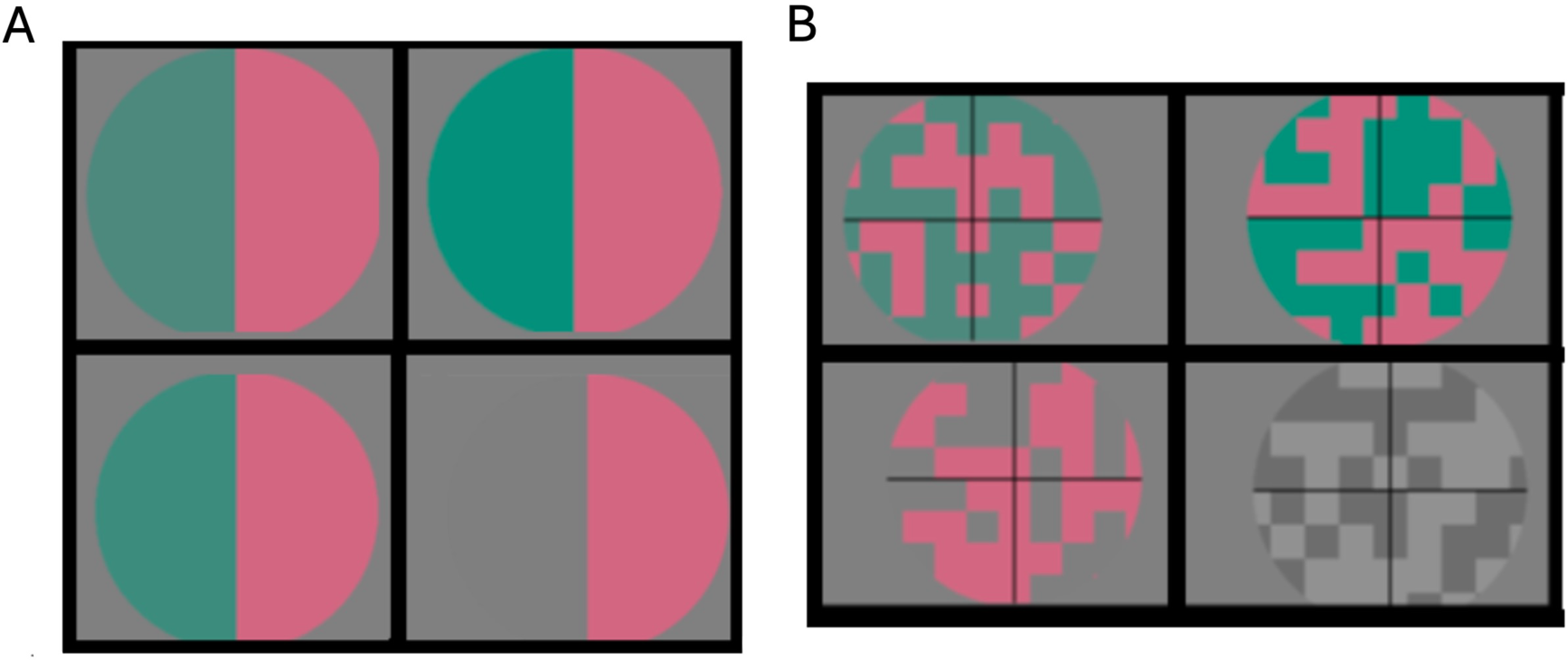
Examples from the psychophysical test and the chromatic stimuli used for achromatic participants. *Left (Panel A*): This panel illustrates four different luminance values of the green semicircle (1.25, 1.075, 1.2, and 1.5, starting from the top left), while the red semicircle remained constant. These luminance values were determined during the psychophysical test and were used to create the experimental stimuli. *Right (Panel B*): This panel shows the stimuli generated based on the green semicircle luminance values. Specifically, the green luminance values are 1.25, 1.075, and 1.5 (starting from the top right). The final example, located in the bottom right, represents the achromatic stimulus used in the experiment.

During both experiments, participants were instructed to fixate on a central point while maintaining attention. For trichromatic and Daltonic participants, the fixation point was a cross (“+”) that changed color non-periodically between red and green. For achromatopsia participants, the fixation point alternated in shape between “+” and “X” to accommodate their inability to perceive color changes. Participants pressed a button whenever the fixation point changed to ensure continuous engagement.

### MRI Data Acquisition

MRI data were acquired on a 3.0T Siemens Prisma scanner with a 64-channel posterior head coil at the Max Planck Institute for Biological Cybernetics, Tübingen, Germany. High-resolution T1-weighted anatomical images were obtained using a magnetization-prepared rapid acquisition gradient echo sequence (ADNI protocol) with the following parameters: voxel size = 1 × 1 × 1 mm³, TR = 2000 ms, TE = 3.06 ms, TI = 1100 ms, flip angle = 9°, and matrix size = 256 × 256. Functional images were acquired using a gradient-echo EPI sequence optimized for occipital cortex activation. Functional scans consisted of 29 slices covering the occipital lobe and cerebellum, with voxel size = 3 × 3 × 3 mm³, TR = 2000 ms, TE = 35 ms, flip angle = 90°, and matrix size = 64 × 64. Each functional run consisted of 181 volumes, and up to five runs were collected per participant. For the retinotopy experiments, we collected at least five repeated scans for each participant to ensure robust and reliable data. Each functional scan consisted of 195 image volumes, with the first three volumes discarded to account for signal stabilization and to minimize noise in the analysis.

### Preprocessing and Analysis

Preprocessing was performed using the MATLAB-based mrVista software (http://white.stanford.edu/software/). Functional images underwent motion correction to mitigate within- and between-run head movements (Gilbert and Wiesel, 1992). Co-registration was applied to align functional images with high-resolution anatomical scans using mutual information-based alignment (Schmid LM, et al. 1996). Gray and white matter were manually segmented using ITK-GRAY, and 3D cortical surfaces were reconstructed to overlay functional activation maps.

For the color/luminance experiment, a General Linear Model (GLM) was employed to estimate voxel-wise activation for each condition, with predictors modeled using a canonical hemodynamic response function (HRF). Two primary contrasts were computed:

1. all conditions vs. baseline, to identify visually responsive voxels, and (2) [color + luminance] vs. luminance, to isolate voxels specifically responsive to chromatic information by subtracting the common luminance component. The proportion of chromatic- and luminance-sensitive voxels within each ROI was calculated and compared across participant groups.

#### Population Receptive Field (pRF) Mapping

To map population receptive fields (pRFs), we used a well-established method inspired by earlier studies (Giannikopoulos DV & Eysel UT, 2006). In simple terms, this approach identifies which part of the visual field activates a specific region of the visual cortex (a voxel). The model assumes a circularly symmetric Gaussian shape for the receptive field, and its center and size (radius) are estimated by comparing actual brain activity (BOLD signals) to predictions generated by the model. These predictions were based on how the moving bar stimuli interacted with the receptive field.

To ensure reliable results, we included only voxels where the model explained at least 10% of the variance in their response. This threshold was determined by measuring the average explained variance (3% ± 2%) in areas unresponsive to visual stimuli and setting the cutoff at three standard deviations above this average. Using this approach, we generated consistent and accurate maps of visual field representation and receptive field sizes in the brain.

The retinotopy data derived from the pRF modeling were also used to define specific retinotopic regions of interest (ROIs) in the visual cortex. These ROIs provided the framework for further analyzing responses to color and luminance stimuli, allowing us to investigate how different visual features are processed in these regions.

#### Visual Field Coverage Maps

To create visual field coverage maps, we identified the locations within the visual field that elicited significant responses from voxels in a given region of interest (ROI) in the visual cortex. This was done by plotting the centers of the population receptive fields (pRFs) for all voxels within the ROI and estimating their sizes using a 2D Gaussian model. The Gaussian was normalized to have a peak amplitude of 1, providing a clear visualization of the coverage.

To enhance the visualization, we generated a color map by assigning each visual field location the maximum normalized pRF amplitude across all overlapping Gaussian pRFs. We also created a non-normalized version of the map (Fig. 2B, second column) where the pRF Gaussians were weighted by their actual response amplitudes. To prevent strong outliers from obscuring weaker responses, the maximum color value was set based on the median pRF amplitude across all voxels in the map.

These reliable pRF measurements and visual field coverage maps were derived using the direct isotropic Gaussian pRF modeling approach (Dumoulin SO &Wandell BA, 2008), ensuring robust and reproducible results. This method allowed us to comprehensively characterize the visual field representation within each ROI.

## Results

### Normal Trichromatic Participants

The results were calculated based on the differences in neural responses across two primary contrasts with a significance threshold of p<0.001. The findings were consistent with theoretical predictions for the control group, confirming expectations based on prior studies. Specifically, as shown in figure 3 (top-right and bottom panels), the hV4 region exhibited the highest sensitivity to chromatic contrasts across all participants, supporting its established role as the primary cortical area responsible for color processing.

**Figure 3:**
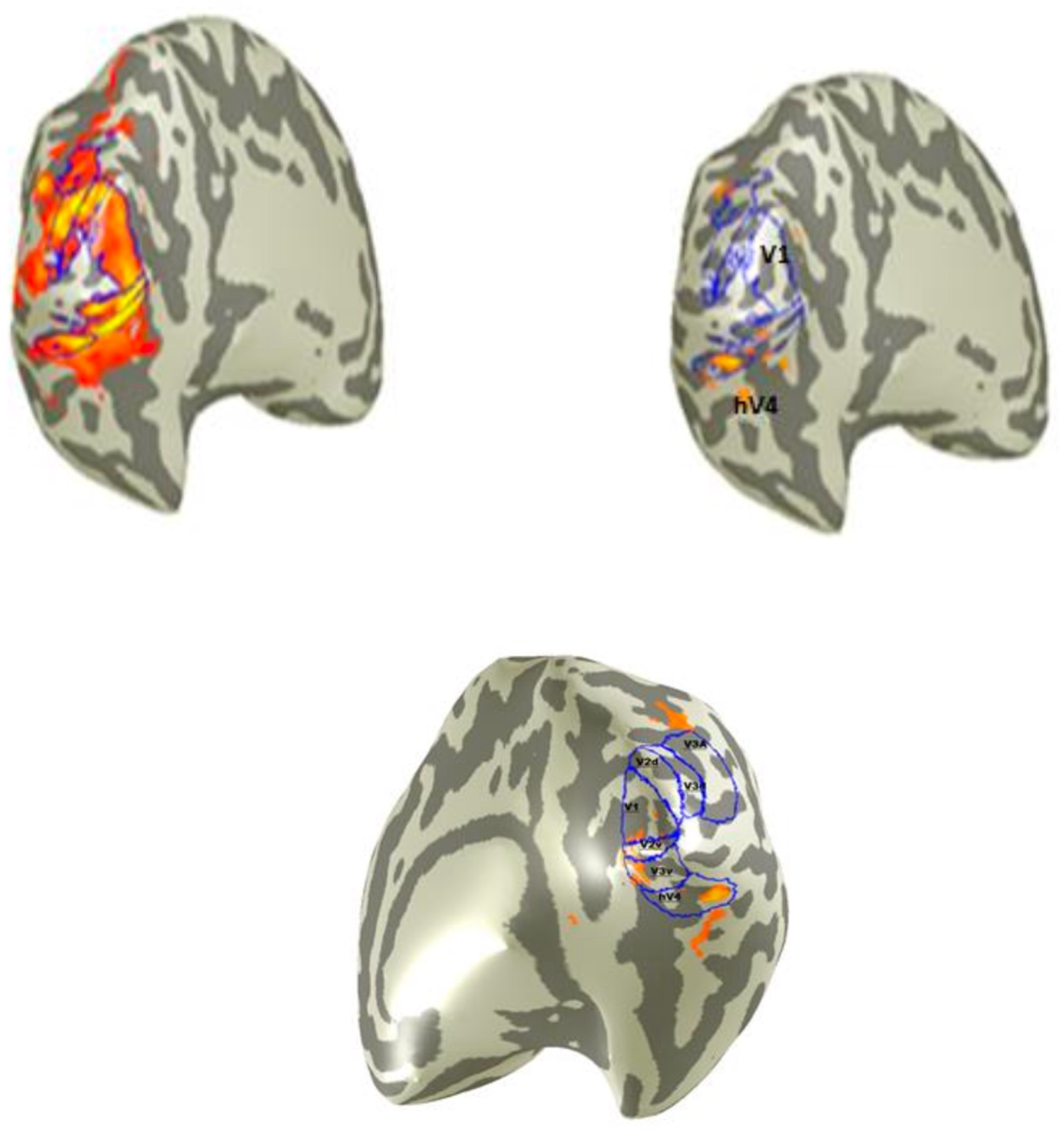
Examples of activations in non-patients. The images display examples of brain activations in non-patient participants. Top left and top right panels come from the same subject, while the bottom panel is from a different subject. The *top left* image shows activations from the comparison of all chromatic conditions versus the intertrial baseline (“All (col. cond.) vs. Intertrial”). The *top right* image applies the contrast “[col+lum.] vs. Luminance” for the same hemisphere of the same subject, highlighting notable differences. Key regions such as V1 and hV4 are indicated. The *bottom image* emphasizes hV4 as the area with the highest sensitivity to color stimuli.

When analyzing the first contrast (all (col. cond.) vs. intertrial), significant activation was observed in the left hemisphere of one participant (Fig. 3 top-left). Adjacent panels revealed substantial differences between cortical regions in response to the [col + lum.] vs. luminance contrast, with the strongest chromatic selectivity localized in hV4. Similar trends were observed in the right hemisphere (Fig. 3, bottom).

In the [col + lum.] vs. luminance contrast, regions such as V3d, V3A, and V2d exhibited minimal to no chromatic selectivity in both hemispheres for 100% of participants (Fig. 4). While V2v and V3v, particularly in the right hemisphere, showed greater activation, their chromatic responses were modest compared to hV4. These findings suggest some chromatic sensitivity in ventral regions but with a dominant preference for luminance processing (Fig. 4 & 5).

**Figure 4:**
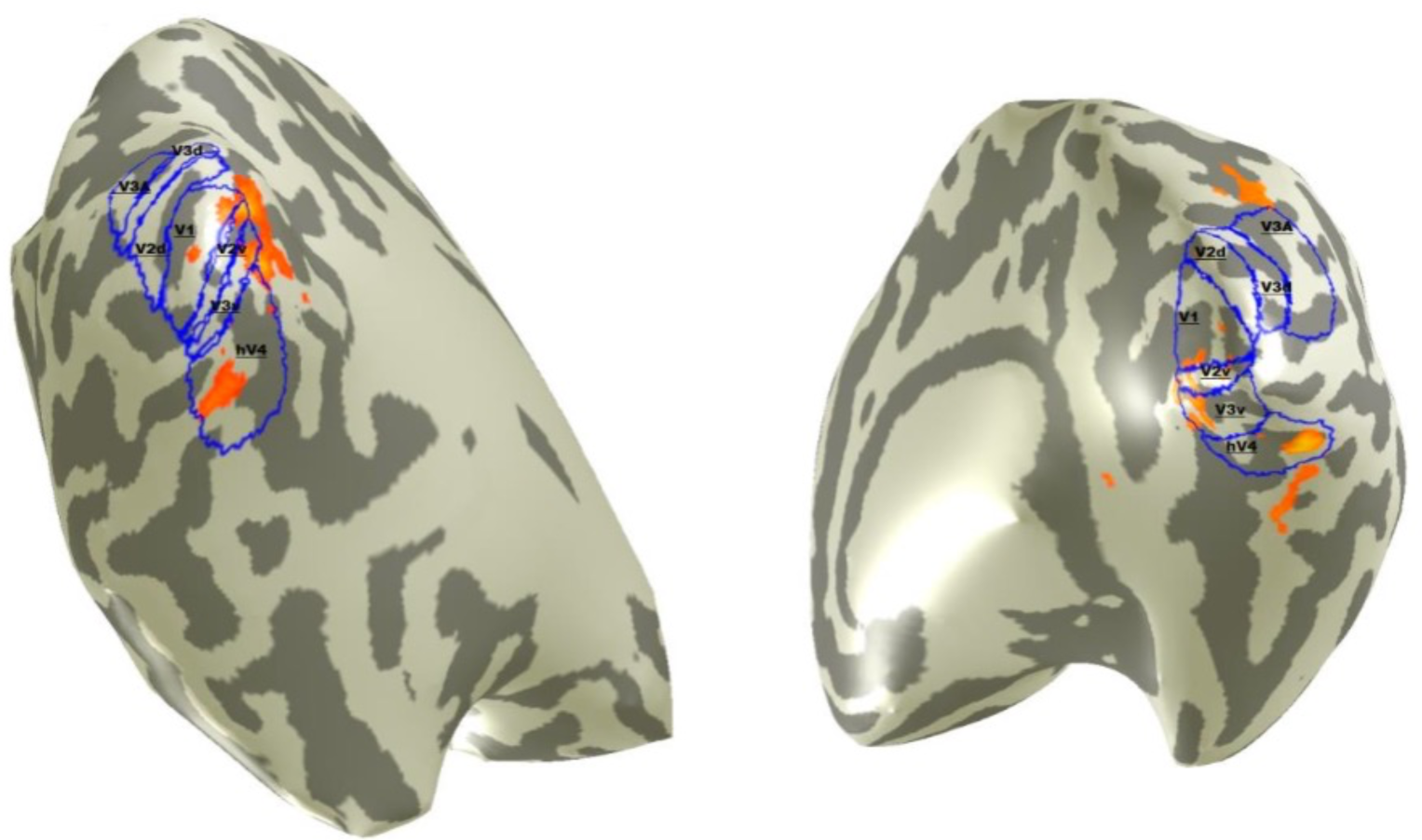
Examples of Activations for “[Color + Luminance] vs. Luminance”. The figure illustrates activation examples in the left and right hemispheres of different participants during the contrast “[Color + Luminance] vs. Luminance,” highlighting regions involved in color processing. In both hemispheres, the highest activation is observed in the hV4 region, followed by V3v and V2v. The activation in V1 is smaller and proportional to its size, while regions such as V2d, V3d, and V3A show minimal to no activation.

**Figure 5:**
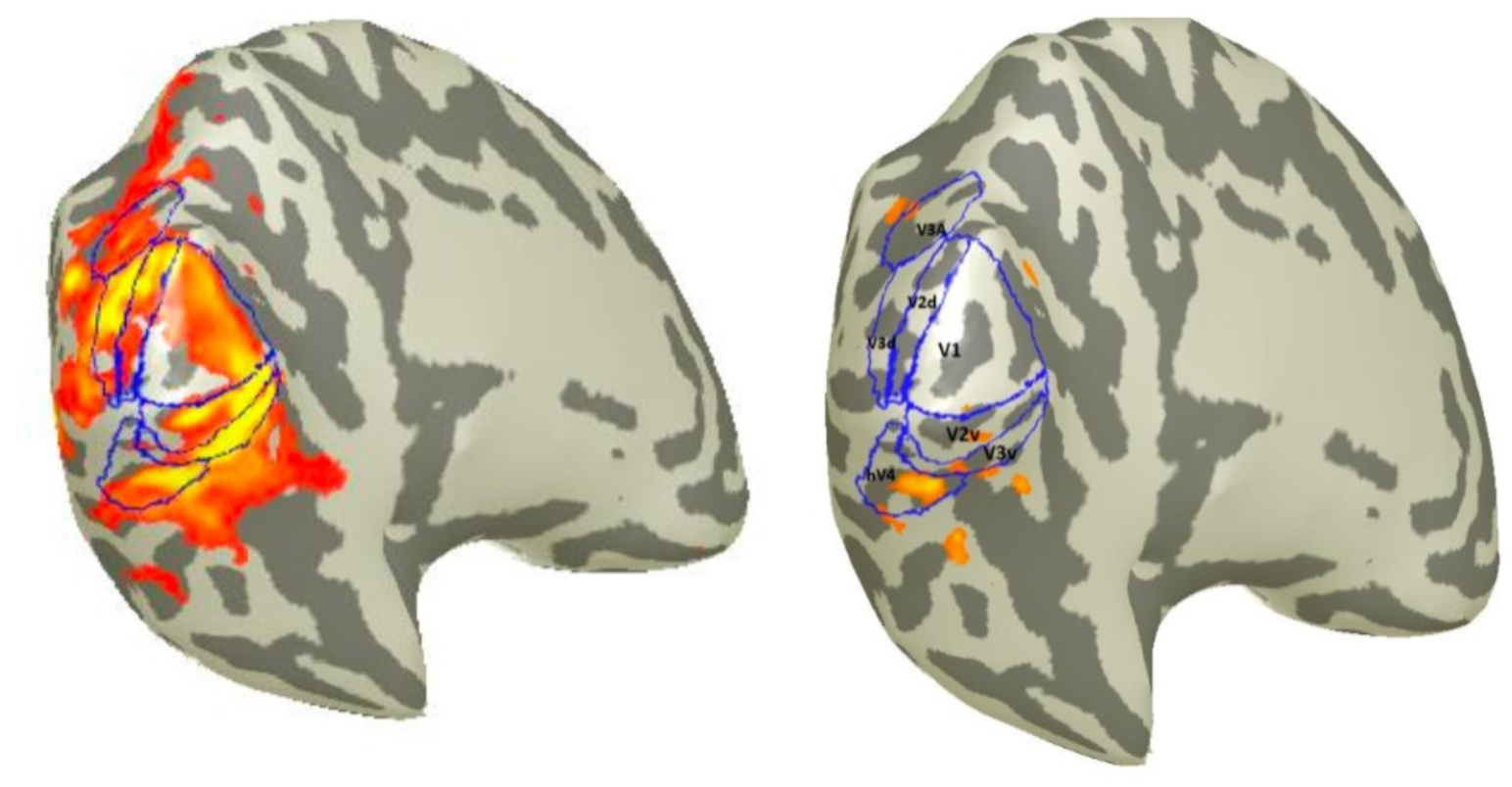
Example of Left Hemisphere Activation in a Non-Patient. The left panel shows the activation induced in the left hemisphere of a non-patient participant during the presentation of all chromatic conditions. The right panel highlights the most color-sensitive regions for the same participant using the contrast “[Color + Luminance] vs. Luminance.” In this case, the dominance of hV4 compared to other regions is clearly evident.

In the primary visual cortex (V1), relatively strong responses to chromatic stimuli ([col + lum.] vs. luminance) were observed, particularly in the right hemisphere. However, the chromatic sensitivity in V1 was significantly lower when compared to its luminance sensitivity, as determined by the all (col. cond. + luminance) vs. intertrial contrast (Fig. 6). Despite this, the robust color responses observed in V1 are consistent with prior findings by Wade et al. (2008).

**Figure 6:**
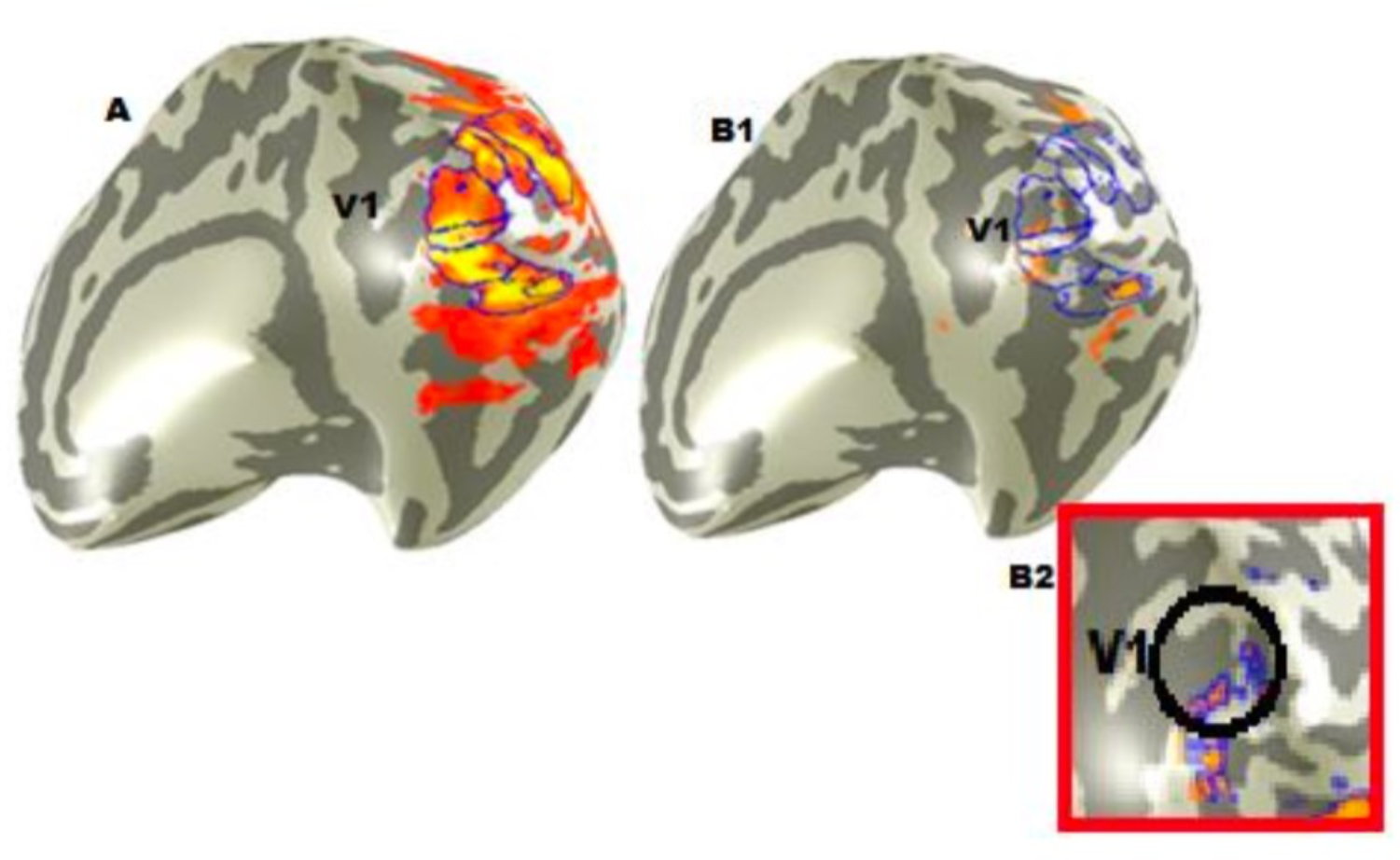
Example of V1 Activation in a Non-Patient. This figure illustrates brain activation in the V1 region of a non-patient participant. Panel A shows the activation induced by the contrast “All (Color Conditions + Luminance) vs. Intertrial,” while Panel B1 shows the activation resulting from the contrast “[Color + Luminance] vs. Luminance.” The difference between the two images is significant, demonstrating that V1 is relatively sensitive to color processing, but to a much lesser extent compared to its sensitivity to luminance. Panel B2 provides a magnified view of the V1 activation area, showing the restricted activation in detail.

**Figure 7:**
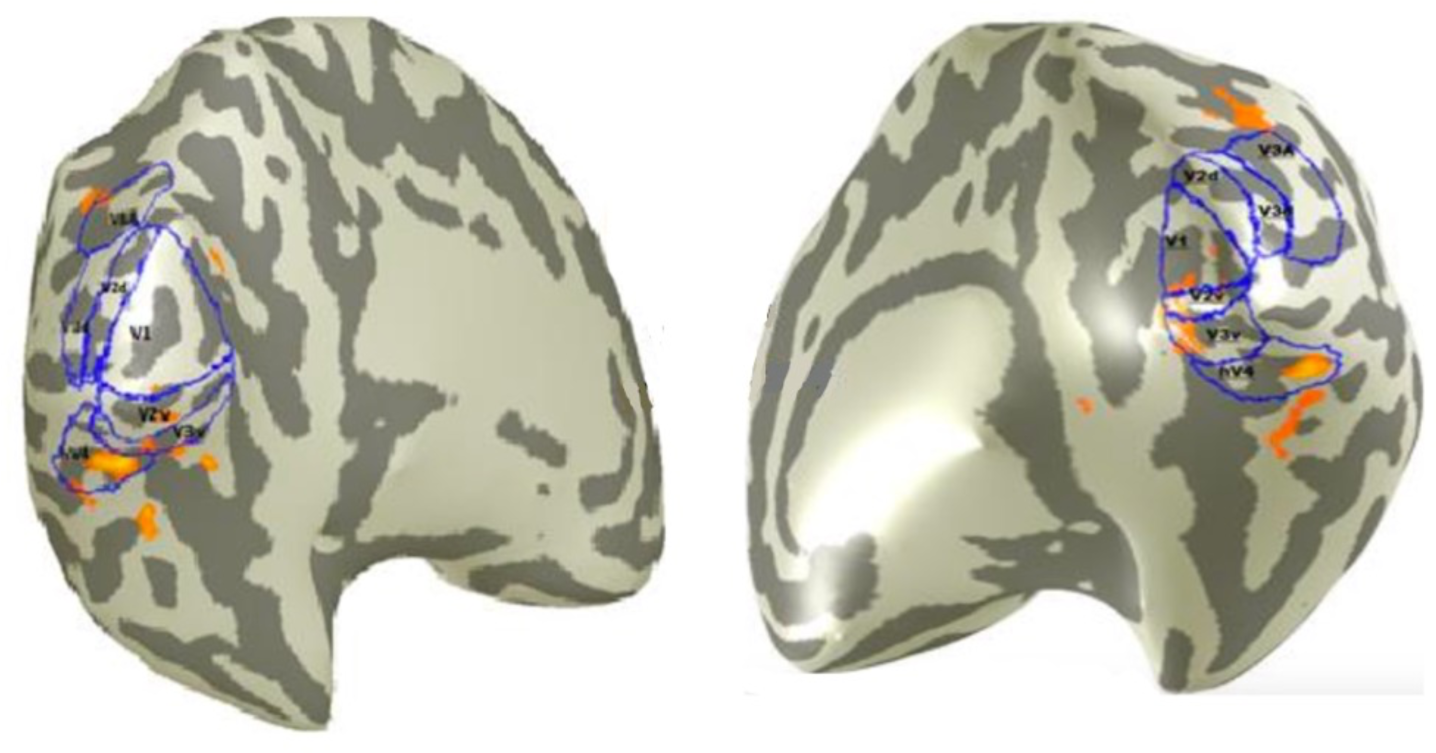
Examples of Activations in Dorsal and Ventral Areas. This figure highlights the differences in activation between dorsal and ventral areas during the application of the contrast “[Color + Luminance] vs. Luminance.” Greater activation is observed in the ventral areas (lower sections of V1, V2v, V3v, and hV4) compared to the dorsal areas (upper sections of V1, V2d, V3d, and V3A).

**Figure 8:**
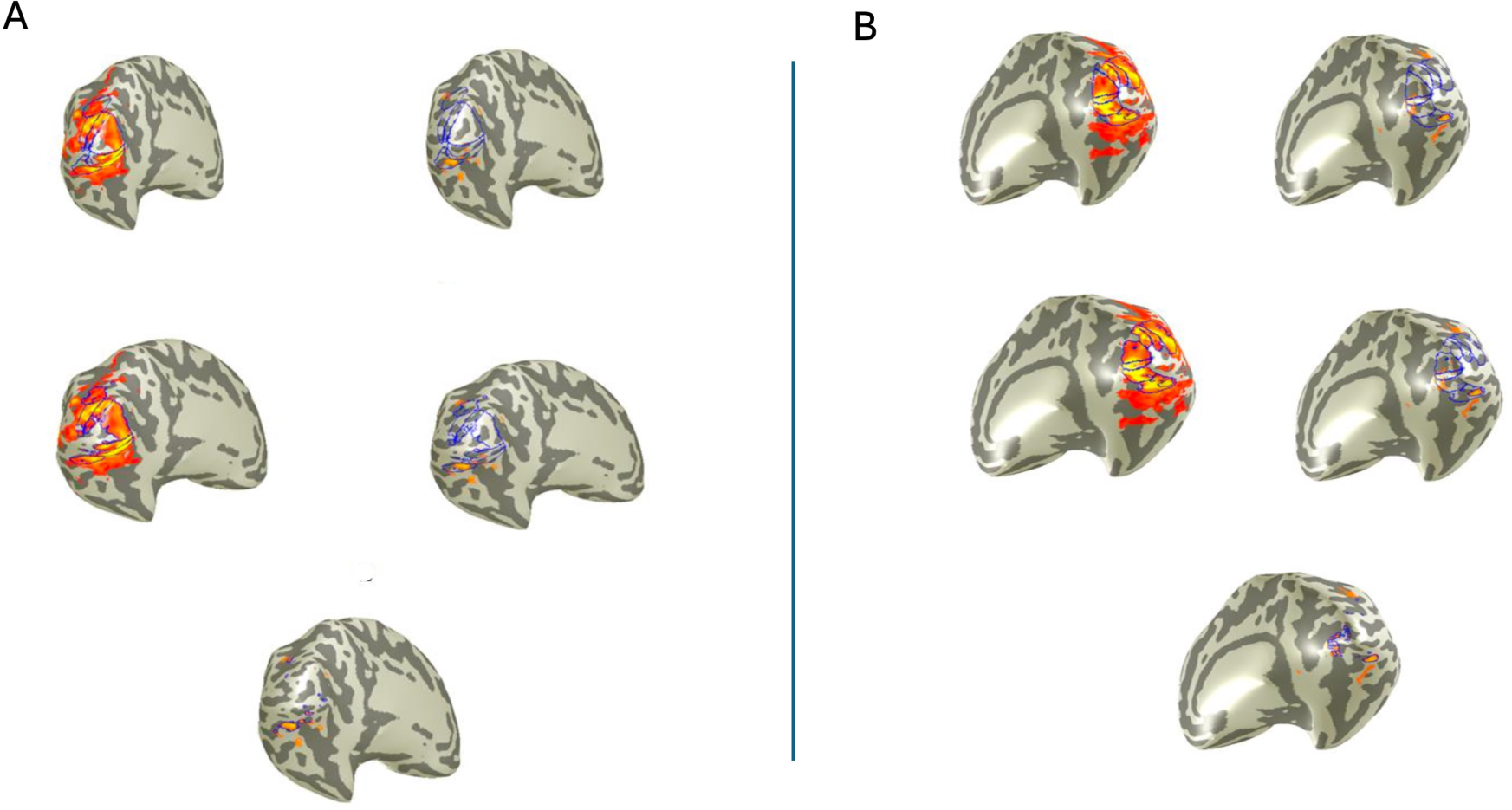
Left and Right Hemisphere Activations in a Non-Patient. A: The *top left and right panels* show activations in the *left hemisphere* for the contrasts *“All (Color Conditions) vs. Intertrial”* and *“[Color + Luminance] vs. Luminance”*, respectively, using non-restricted ROIs. The *middle panels*display the same contrasts but with restricted ROIs. In the *middle right panel*, the restricted ROIs are shown to visually emphasize the differences. The quantitative distinction between these contrasts is further highlighted in the *bottom panel*, where the restrictions are reapplied, revealing a clearer difference. Across all panels, the *hV4 region* consistently appears as the most influenced by color processing, confirming its role in color sensitivity. B: The same analyses are applied to the *right hemisphere*, showing a similar pattern of activations.

**Figure 9:**
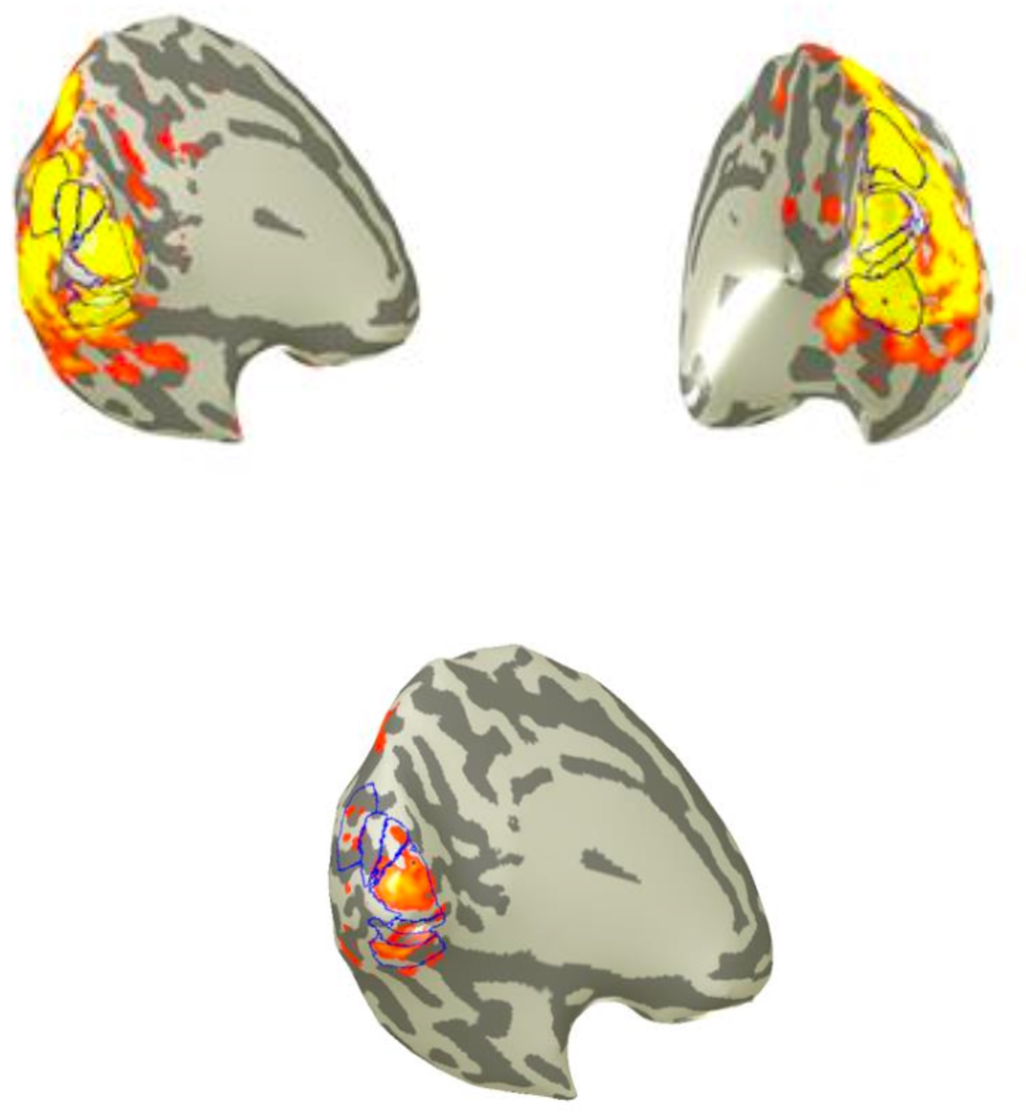
Brain activations for two contrasts in both hemispheres of a participant. *Top*: Activations in the *left* and *right hemispheres* are shown for the contrast *“All (Color Conditions) vs. Intertrial.”*Widespread activation is observed across visual regions, reflecting the combined influence of chromatic and luminance signals. *Bottom*: Activations in the *left hemisphere* of the same participant for the contrast *“[Color + Luminance] vs. Luminance.”*Here, activation remains visible, particularly in areas like *hV4*, which are typically associated with color processing. This suggests that the participant’s brain responds to chromatic information even when luminance is removed. Luminance enhances overall visual activation, but chromatic signals alone are still sufficient to engage color-sensitive regions, such as *hV4*.

Additionally, a distinct ventral-dorsal gradient in chromatic responses was noted, with ventral areas showing stronger chromatic activation compared to dorsal regions. This observation aligns with the findings of Bartels and Zeki (2000) and Wade et al. (2008) (Figure 0.5). The amplitude of responses to chromatic contrasts in hV4 was notably higher than those in neighboring regions such as V3v and surpassed responses across all other visual cortical areas (Wade et al., 2008). By contrast, hV4 exhibited no specific preference for luminance stimuli when compared to adjacent regions under the luminance-only condition.

### Daltonic Participant

For the Daltonic participant, results from the all (col. cond. + luminance) vs. intertrial contrast (Fig.10A-left) revealed robust activation across early and mid-level visual regions, including V1, V2, V3v, and hV4. This pattern indicates that visual stimuli containing both chromatic and luminance components were effectively processed, driven primarily by luminance differences. The extent and distribution of activation suggest that luminance-based visual pathways remain intact and functional, even in the absence of red-green chromatic processing.

In contrast, the [col + lum.] vs. luminance condition (Fig. 10A-right) showed no significant activation in any visual region, including hV4, which is typically specialized for chromatic processing. This absence of activity reflects the participant’s inability to process chromatic information independently of luminance cues, a limitation consistent with their L-cone deficiency and dichromatic vision. Similar findings were observed in the right hemisphere (Fig. 10B), where robust activation in all (col. cond. + luminance) vs. intertrial (Fig. 10B-left) contrasted sharply with the absence of responses in [col + lum.] vs. luminance (Fig. 10B-right).

**Figure 10:**
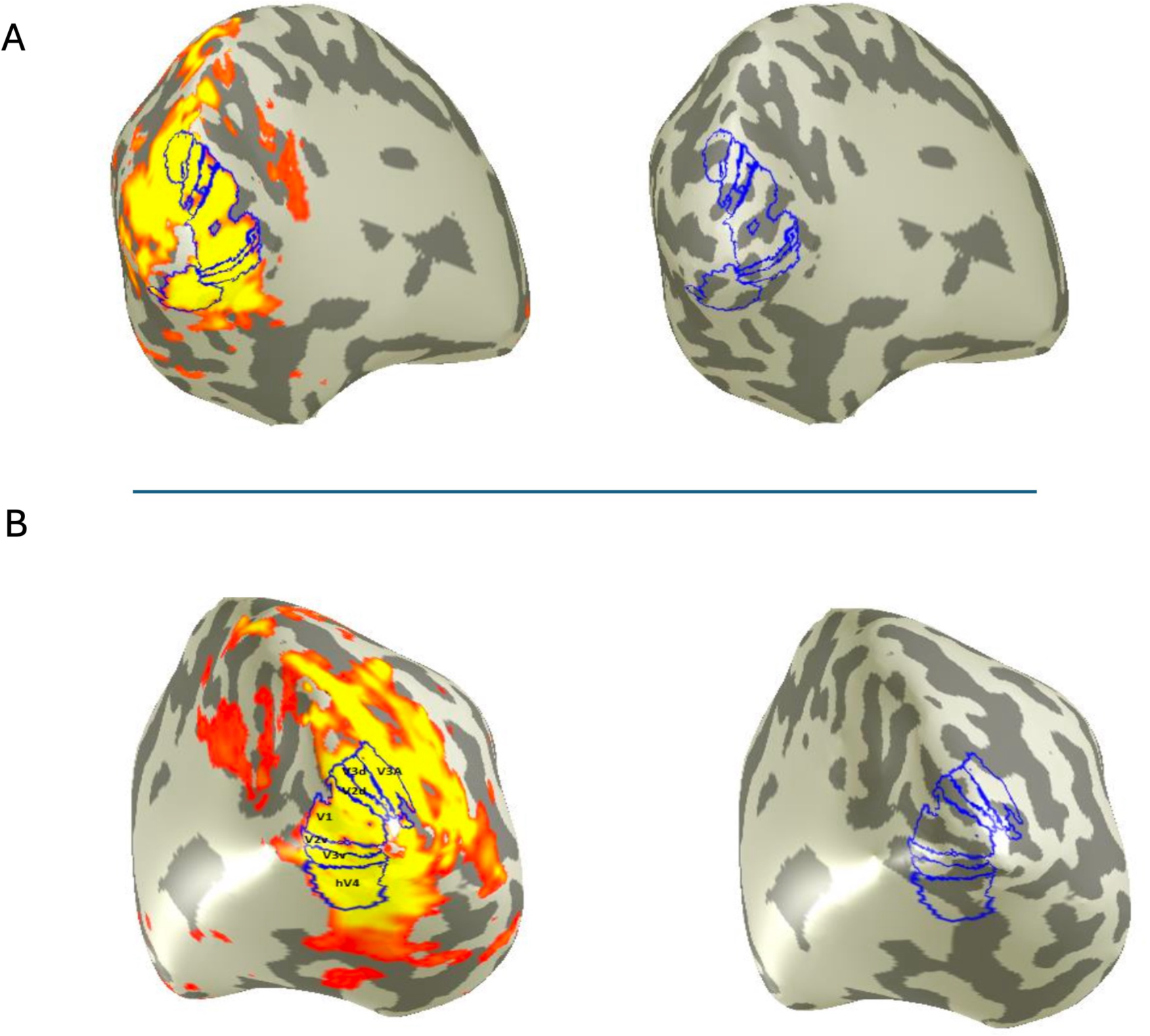
Brain Activations in the Left and Right Hemispheres of a Daltonic Participant. *A*: Brain activations in the *left hemisphere* of a participant with red-green color vision deficiency (daltonism). The *left image* shows activation for the contrast *“All (Color Conditions + Luminance) vs. Intertrial”*, reflecting responses driven by luminance information. In contrast, the *right image*, representing the contrast *“[Color + Luminance] vs. Luminance”*, shows no significant activation. This lack of response suggests an inability to process isolated chromatic signals, consistent with impaired *L*- and *M*-cone function. *B*: The same contrasts are displayed for the *right hemisphere* of the same participant. The *left image* again shows activation for *“All (Color Conditions + Luminance) vs. Intertrial”*, highlighting the role of luminance in driving responses. However, the *right image* confirms the absence of activation for the contrast *“[Color + Luminance] vs. Luminance”*, reinforcing the participant’s inability to process purely chromatic stimuli. While luminance signals can still elicit robust activations, purely chromatic signals fail to engage color-sensitive areas, such as *hV4*, in participants with red-green color vision deficiency.

The results provide clear evidence that the participant relies exclusively on luminance cues to perceive and differentiate visual stimuli. The activation observed in ventral regions, such as V3v and hV4, during the all (col. cond. + luminance) vs. intertrial contrast underscores the strength of luminance-driven pathways. Notably, the ventral dominance of activation suggests that these regions, while typically chromatic-sensitive, may still process detailed luminance-based attributes in the absence of chromatic information.

### Achromatopsia Participants

For the achromatopsia participants, the all (col. cond. + luminance) vs. intertrial contrast (Fig.11A & B-left) revealed modest activation in early visual areas, such as V1 and V2. These regions responded to the luminance cues embedded in the stimuli, enabling basic visual processing through rod-driven mechanisms. The activation was spatially restricted and weaker compared to that observed in trichromatic participants, particularly in mid-level regions like V3v and hV4, reflecting the functional limitations of rod-mediated vision.

**Figure 11:**
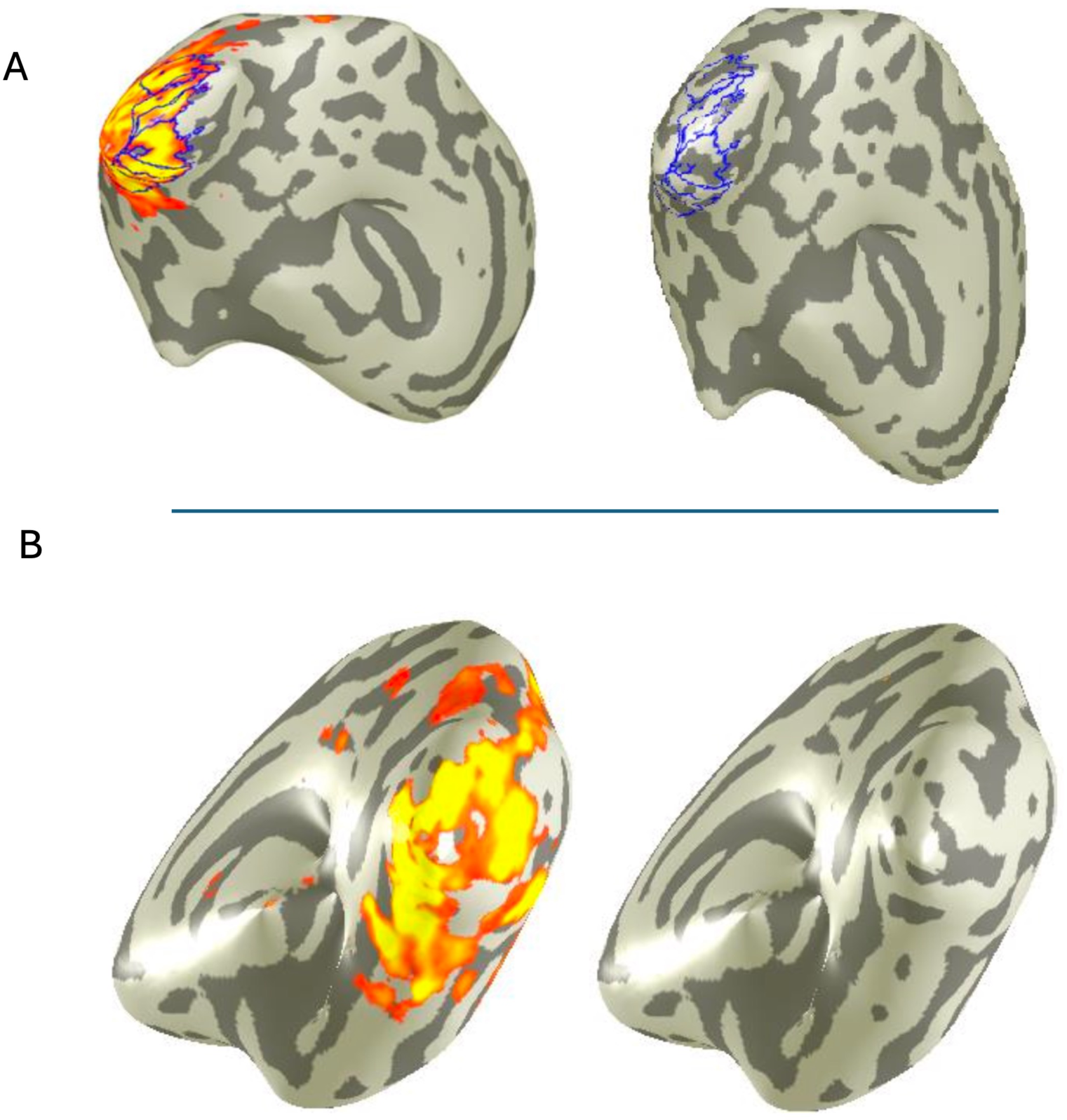
Brain Activations in the Left and Right Hemispheres of a CNGA3-Achromatopsia Participant. A: Brain activations in the left hemisphere of a participant with CNGA3-achromatopsia. In the left image, the contrast “*All (Color Conditions + Luminance) vs. Intertrial*” shows clear activation, driven by luminance signals, likely mediated by rod pathways. In the right image, the contrast *“[Color + Luminance] vs. Luminance*” isolates the chromatic signal but shows no significant activation. This confirms the participant’s inability to process color information due to non-functional cones. B: Brain activations in the right hemisphere of the same participant. The left image again shows activation for the contrast “*All (Color Conditions + Luminance) vs. Intertrial*”, consistent with a luminance-driven response. The right image, corresponding to the contrast *“[Color + Luminance] vs. Luminance*”, reveals no activation, further demonstrating the absence of chromatic sensitivity. These results highlight that visual responses in CNGA3-achromatopsia participants are dominated by rod-mediated luminance processing. The lack of cone functionality prevents chromatic information from being processed, resulting in no measurable activation in color-selective regions like hV4.

In contrast, the [col + lum.] vs. luminance condition (Fig. 11 A&B - right) showed no significant activation in any cortical region, including areas such as hV4, which is typically specialized for chromatic processing. This absence of activation underscores the participants’ complete reliance on luminance-based pathways and the lack of functional cone-mediated input. Notably, these results were consistent across both hemispheres, highlighting the bilateral nature of the visual deficits in achromatopsia.

## Discussion

This study investigated the cortical processing of chromatic and luminance stimuli in participants with normal trichromatic vision, a Daltonic participant, and two participants with complete achromatopsia. Using functional magnetic resonance imaging (fMRI), we aimed to explore the activations caused by chromatic and luminance stimuli in the visual cortex of healthy participants, identify regions responsible for color and luminance processing, and compare these activations to those observed in individuals with color vision deficiencies.

The stimuli consisted of four conditions: (1) Luminance Contrast Only (L+M+S), where squares varied in brightness with random achromatic contrasts up to 24%, modulating all three cone types equally; (2) Chromatic Contrast Only (Red-Green; L-M), where chromatic contrasts of up to 6% were achieved by modulating L (red-sensitive) and M (green-sensitive) cones in opposite directions while maintaining constant S cone excitation and overall luminance; (3) Chromatic Contrast Only (Blue-Yellow; L+M-S), where L+M cones were modulated relative to the S cone, maintaining constant overall luminance; and (4) Combined Luminance and Chromatic Contrast (L+M+S + L-M), combining brightness and hue variations with simultaneous modulation of luminance and chromatic contrasts.

Data were collected from seven participants: four with normal trichromatic vision, one Daltonic participant, and two participants with complete achromatopsia. To assess overall cortical activation, we performed a contrast (“all col. cond. vs. intertrial”, p=0.001), comparing all chromatic and luminance conditions against a baseline intertrial condition. To examine activation driven specifically by color, we conducted a second contrast (“[col + lum.] vs. luminance”, p=0.001), comparing the chromatic stimuli with added luminance to the luminance-only condition.

Our findings confirm that color processing in trichromatic participants is primarily mediated by ventral visual stream regions, including V1, V2v, V3v, and hV4, with hV4 showing the strongest chromatic-specific activation. These results align with previous studies (Engel et al., 1997; Bartels & Zeki, 2000; Conway & Tsao, 2006; McKeefry & Zeki, 1997; Barbur & Spang, 2008; Leeuwen et al., 2014) and emphasize the specialization of hV4 in processing chromatic information. V1 exhibited significant activation for red-green stimuli, consistent with Engel et al. (1997), though its activation remained relatively lower compared to luminance stimuli. Dorsal regions, such as V3d and V3A, showed negligible activation, consistent with their limited role in chromatic processing (Adams & Zeki, 2001; Felleman & Van Essen, 1987; Tsao et al., 2003; Wade et al., 2002). In contrast, V3v and V2v displayed significant activation compared to V2d, consistent with the ventral stream’s role in chromatic processing (Conway & Tsao, 2006; Wade et al., 2002).

For the Daltonic participant, activation patterns were similar to trichromatic participants in the “all col. cond. vs. intertrial” contrast (p=0.001), reflecting intact luminance processing mechanisms. However, the absence of activation in the “[col + lum.] vs. luminance” contrast (p=0.001) underscores the participant’s reliance on luminance cues for distinguishing red-green stimuli. This finding is consistent with metamerism, wherein the absence of M-cones results in the misclassification of red-green colors based on luminance differences. Metamerism expands the perceptual similarity of colors, causing confusion in red-green discrimination tasks. These results also highlight the challenges faced by dichromatic individuals in tasks requiring fine chromatic discrimination (Engel et al., 1997; Deeb, 2005).

Achromatopsia participants exhibited a unique activation profile. Responses in the “all col. cond. vs. intertrial” contrast were limited to early visual areas (V1, V2), with no significant activation in higher-order regions such as hV4. This restricted activation pattern reflects the reliance on rod-mediated luminance processing, as the absence of functional cones precludes chromatic input. The complete lack of activation in the “[col + lum.] vs. luminance” contrast further confirms the inability of achromatopsia participants to process chromatic stimuli. Interestingly, overall cortical activation for luminance stimuli was markedly lower in achromatopsia participants compared to other groups, likely reflecting the reduced capacity of rod-dominated pathways to encode luminance information compared to combined rod- and cone-mediated inputs (Carlson, 2013). This diminished activation in the occipital lobe highlights the challenges of rod-only vision.

### Clinical Implications and Future Research Directions

Our findings have significant implications for understanding cortical plasticity and the potential for visual rehabilitation in individuals with color vision deficiencies. For the achromatopsia participants, this study provides baseline data for future research on gene therapy aimed at restoring cone functionality. By comparing pre- and post-treatment cortical responses, we aim to assess the extent of functional recovery in chromatic-sensitive regions such as hV4.

Future studies should address the limitations of this research. Increasing the sample size, particularly in color-deficient groups, will enhance the statistical power and generalizability of the findings. Moreover, age- and sex-matched participant groups would help control for potential confounding variables. Additionally, behavioral data (e.g., reaction times, accuracy) should be incorporated to correlate neural activation with task performance, particularly for the Daltonic participant, where luminance-based discrimination may influence the results.

Finally, integrating advanced analytical approaches such as functional connectivity analyses could provide insights into the interactions between visual regions during chromatic and luminance processing. These methods could also help identify compensatory mechanisms in individuals with color vision deficiencies.

### Contribution to the Field

This studymakes significant contributions to the understanding of visual processing in both trichromatic individuals and those with color vision deficiencies. By establishing baseline cortical data for achromatopsia patients undergoing gene therapy, the research provides a foundation for future longitudinal comparisons of cortical recovery and plasticity. Specifically, it highlights the limited role of rod-dominated pathways in luminance processing and their reduced efficacy compared to cone-mediated inputs. For the Daltonic participant, the findings emphasize the reliance on luminance cues for chromatic discrimination and provide robust evidence of metamerism using advanced fMRI contrasts.

The study also refines our understanding of chromatic processing by confirming the dominance of ventral visual stream areas, such as hV4, in trichromatic individuals, while documenting negligible activation in dorsal regions. These findings align with previous research but offer a more detailed perspective on the segregation of chromatic and luminance processing pathways. Methodologically, the research introduces a tailored psychophysical calibration protocol for achromatopsia participants, ensuring isoluminance during stimulus presentation, which enhances the precision of neural activation measurements.

Together, these contributions advance the field by providing new insights into the cortical mechanisms underlying color vision deficiencies, offering a framework for future studies on visual rehabilitation, cortical plasticity, and the effects of gene therapy.

## Author Contributions

This study was conducted in collaboration with the Max Planck Institute for Biological Cybernetics, Tübingen, Germany and under the supervision of Dr.Georgios A.Keliris. The research focused on investigating cortical responses to chromatic and luminance stimuli in trichromatic individuals and those with color vision deficiencies. AR designed and built the experiments, recruited participants, collected the data, conducted all analyses, and wrote the manuscript. AR also developed the psychophysical calibration methods to tailor the experiment for achromatopsia participants and ensured the validity and precision of the experimental design.

## Data Availability

All data produced in the present study are available upon reasonable request to the authors

